# Do Cognitive Symptoms Improve When Gabapentin and Dihydropyridine Calcium Channel Blocker Therapy Are Stopped? A Self-Controlled Case Series

**DOI:** 10.64898/2026.03.30.26349787

**Authors:** James W. Green, Joshua Kaplan, Branimir Ljubic, Michael Schulewski, Suril Gohel, Sharon Sanz Simon, Luciana Mascarenhas Fonseca, Michal Schnaider Beeri, Laura Byham-Gray, Barbara Tafuto

## Abstract

**Objective:** Older adults presenting with cognitive complaints are frequently taking medication regimens that include gabapentinoids and antihypertensive calcium channel blockers, yet whether such symptoms improve after discontinuation has been poorly characterized. We examined whether cognitive symptoms associated with concomitant gabapentin and dihydropyridine calcium channel blocker (DHP-CCB) use decline following discontinuation, with secondary analyses of falls and encephalopathy and pre-specified evaluation of dihydropyridine-class specificity and chronic kidney disease (CKD) amplification.

**Methods:** We conducted a self-controlled case series in two independent cohorts: the NIH All of Us Research Program Controlled Tier (primary cohort, N=8,853) and the Rutgers Clinical Research Data Warehouse (CRDW; triangulation cohort, N=2,404). Within each patient, event rates during the final equal-duration window of concomitant gabapentin–DHP-CCB use were compared with an equal-duration window following discontinuation. Primary outcomes were cognitive symptoms and a composite cognitive endpoint; secondary outcomes were falls, encephalopathy, and delirium. Pre-specified analyses included a non-DHP CCB negative control, CKD stratification, and lag-period sensitivity analyses. An exploratory analysis in patients with established dementia (N=246) is reported in the Supplementary Material.

**Results:** Cognitive symptom rates were higher during concomitant gabapentin–DHP-CCB use than after discontinuation in both cohorts. The decline was most pronounced in patients who stopped both medications, where the All of Us rate ratio was 1.45 (95% CI 1.35–1.57, p<0.001) and the CRDW estimate was closely concordant at 1.46 (95% CI 1.09–1.94, p=0.012) — strengthening to 1.74 (95% CI 1.22–2.50, p=0.003) in adults aged ≥65, the population most relevant to deprescribing. Secondary outcomes (falls, encephalopathy) were directionally consistent in the larger All of Us cohort, the non-DHP CCB negative control was null for cognitive symptoms, and CKD amplified effects across primary outcomes.

**Conclusions:** Cognitive symptoms associated with concomitant gabapentin–DHP-CCB use declined after discontinuation, with the largest improvements among patients who stopped both medications and a closely concordant signal across two independent cohorts. These findings support medication review and structured deprescribing assessment for older adults presenting with cognitive complaints while taking this combination, and identify cognitive symptoms in this population as a potentially reversible, medication-related contribution to clinical presentation.

## INTRODUCTION

Clinicians caring for older adults frequently encounter patients whose new or worsening cognitive symptoms — confusion, memory complaints, disorientation, or unexplained falls — coincide with the medications they are taking, yet whether these symptoms improve after discontinuation is poorly characterized for many commonly co-prescribed drug combinations. Drug-induced cognitive impairment is estimated to account for 2.7–10% of dementia diagnoses, but reversibility upon discontinuation remains inadequately documented even for well-studied classes such as anticholinergics.^9,10^ As Campbell and Boustani have noted, demonstrating an association between a medication and cognitive impairment does not establish whether stopping the drug restores function — and that question is what determines whether deprescribing is a reasonable clinical step.^11^

One drug pairing in which this evidence gap is clinically pressing is the combination of gabapentin and dihydropyridine calcium channel blockers (DHP-CCBs). Gabapentin is dispensed to approximately 15.5 million patients annually in the United States and is commonly co-prescribed with antihypertensive DHP-CCBs such as amlodipine in older adults.^1,2^ Preliminary work from our group reported that DHP-CCBs may amplify gabapentin-associated dementia diagnosis risk in a new-user cohort, with the association specific to dihydropyridines and not observed with verapamil or diltiazem.^3^ Both drug classes act on neuronal calcium signaling — gabapentin via the α2δ-1 subunit of voltage-gated calcium channels, and DHP-CCBs via L-type calcium channels — providing a biologically plausible basis for a pharmacodynamic interaction.^4–8^ However, that prior work characterized association, not reversibility.

Whether cognitive symptoms associated with this combination decline after discontinuation has direct relevance to clinical practice — to medication review in older adults with cognitive complaints, to deprescribing decisions when a contributing medication is suspected, and to the broader question of how much medication-related cognitive impairment in geriatric populations may be amenable to drug withdrawal. A pharmacodynamic interaction at calcium-dependent synaptic processes — as opposed to irreversible neuronal injury — would be expected to attenuate as drug levels normalize, but this expectation requires empirical evaluation in real-world data.

We therefore evaluated whether cognitive symptoms associated with concomitant gabapentin and DHP-CCB use decline following discontinuation, with the largest decline expected in patients who stop both medications. We tested this question across two independent data sources — the NIH All of Us Research Program Controlled Tier (primary cohort, N=8,853) and the Rutgers Clinical Research Data Warehouse (CRDW; triangulation cohort, N=2,404) — using a within-person comparison of event rates before and after discontinuation. Cognitive symptoms and a composite cognitive endpoint were the primary outcomes; falls, encephalopathy, and delirium were examined as supportive secondary outcomes. Pre-specified secondary analyses examined dihydropyridine-class specificity (versus a non-DHP CCB negative control) and CKD amplification, given that gabapentin clearance is renally dependent. An exploratory analysis in patients with established dementia (N=246) is reported in the Supplementary Material.

## METHODS

### Data Sources

#### NIH All of Us Research Program

The primary analytic cohort was drawn from the All of Us Controlled Tier Dataset (C2024Q3R9; N=633,547 enrolled participants), accessed through the Researcher Workbench.^12^ The Controlled Tier provides standardized OMOP Common Data Model data with SNOMED CT vocabulary. All analyses adhered to All of Us data use policies. Access to the Controlled Tier was provided under the All of Us IRB-approved protocol; the study was determined to be non-human subjects research per 45 CFR 46.102(d).

#### Rutgers Clinical Research Data Warehouse (CRDW)

The single-center triangulation cohort was drawn from the Rutgers Clinical Research Data Warehouse (CRDW)^13,14^, a centralized research repository that integrates electronic health record (EHR) data from Rutgers-affiliated clinical sites across New Jersey, including inpatient and outpatient encounters, diagnoses (International Classification of Diseases, Tenth Revision [ICD-10] codes), medication orders, laboratory results, procedures, and demographic information. This analysis used a hypertension-focused research extract from the CRDW comprising 541,539 patients with documented encounter, medication, and comorbidity data from 2018 to 2024, serving the Rutgers University health system in northern New Jersey.

### Self-Controlled Case Series Design

#### Study Population

In simple terms, each patient served as their own control: event rates while concomitantly taking gabapentin and a DHP-CCB were compared with event rates after one or both medications were stopped. In each cohort, we identified patients with documented concomitant use of gabapentin and a DHP-CCB (amlodipine, nifedipine, felodipine, or nicardipine) for at least 90 days who subsequently discontinued one or both medications, and applied a self-controlled case series (SCCS)^15^ design comparing event rates within the same individual. Discontinuation was defined as no prescription fill within 90 days of the last recorded fill, with at least 90 days of subsequent follow-up thereafter.

#### Exposure Periods

For each patient, we defined two equal-length observation windows anchored to the discontinuation date: a PRE window covering concomitant gabapentin–DHP-CCB use immediately before discontinuation, and a POST window of the same length beginning the day discontinuation took effect. Window length was capped at 180 days.

#### Outcomes

Primary outcomes were (1) cognitive symptoms (ICD-10 R41.0–R41.3, R41.81–R41.82 — confusion, disorientation, amnesia, cognitive decline, and altered mental status, representing clinician-documented acute or subacute cognitive complaints rather than a dementia diagnosis), and (2) a composite cognitive endpoint combining cognitive symptoms with delirium (F05), encephalopathy (G92, G93.4), and incident dementia (F01–F03, G30, G31.0, G31.1, G31.83). Secondary outcomes were falls (W00–W19, R29.6) and the individual components of the composite (delirium, encephalopathy, incident dementia) reported separately. SNOMED CT concept IDs used in the All of Us cohort are listed in the Supplementary Material.

#### Design Rationale and Bias Considerations

The 90-day discontinuation threshold and 180-day matched window follow standard pharmacoepidemiology conventions for these drug classes. To address potential protopathic bias — discontinuation triggered by an emerging adverse event — we pre-specified lagged POST windows of 30 and 60 days (described below). Window-selection rationale and the within-person sign test are detailed in the Supplementary Material.

#### Statistical Analysis

Event rates per 1,000 person-days were compared using Poisson rate ratios (RR) with 95% confidence intervals; values greater than 1.0 indicate higher event rates during combination use than after discontinuation. Pre-specified stratified analyses by discontinuation pattern tested whether benefit was greater from stopping both drugs.

#### Protopathic Bias Sensitivity Analysis

The primary analysis was repeated after shifting the POST window forward by 30 and 60 days, with the same approach applied to the stopped-both cognitive symptoms subgroup in the CRDW cohort. Under pharmacological washout, the rate ratio should strengthen as the immediate post-discontinuation period is excluded.

### Medication Exposure Ascertainment

Medication exposure in the CRDW cohort was identified from the CRDW prescription file (Hypertension Medications 20250417) using a generic-name-first matching strategy. For each prescription record, we matched on the standardized generic name (SIMPLE_GENERIC) when present, falling back to the full medication name (MEDICATION_NAME) when SIMPLE_GENERIC was missing. To restrict the analysis to sustained outpatient oral therapy, we excluded prescription records with ACTIVE_ORDER_STATUS = ‘Discontinued Medication’ and excluded intravenous formulations identified by route (MED_ADMIN_ROUTE containing ‘intravenous,’ ‘injection,’ ‘iv push,’ ‘iv bolus,’ or ‘iv drip’) or by product-name pattern (e.g., ‘…intravenous solution,’ ‘…in 0.9% sodium chloride,’ ‘…mg/mL intravenous’). Drug-class definitions (DHP-CCB, non-DHP-CCB, ACE-inhibitor, ARB) used the standardized generic name list specified in the Supplementary Methods. Ascertainment was performed at the prescription-record level prior to cohort-level eligibility filtering. All of Us medication exposures were ascertained from the OMOP Common Data Model drug_exposure table using RxNorm ingredient identifiers (gabapentin: 1310756; DHP-CCB ingredients: amlodipine 1318137, felodipine 1332418, nifedipine 1353776), restricted to ambulatory dispensing records.

### Cross-Cohort SCCS Implementation

The SCCS protocol was applied identically in both the All of Us Controlled Tier and the CRDW. A non-DHP CCB negative control cohort (gabapentin + verapamil or diltiazem; N=2,011 in All of Us) tested whether reversal signals were specific to DHP-class blockade. Pre-specified CKD stratification (any CKD diagnosis at or before combination start; All of Us N=1,692 CKD, N=7,161 non-CKD) evaluated pharmacokinetic amplification, given that gabapentin clearance is renal.

### Established Dementia Cohort (Supplementary Material)

An exploratory analysis in patients with documented prior dementia who initiated a gabapentinoid (gabapentin or pregabalin) is reported in the Supplementary Material. The supplementary analysis uses a broader exposure definition than the primary SCCS (gabapentinoid versus gabapentin only) to maximize statistical power in this small subpopulation. Under corrected medication ascertainment, the analytic Cox cohort comprised 246 patients (101 DHP-CCB-exposed and 145 no-CCB); an additional 8 non-DHP CCB patients were treated as a descriptive arm and not included in formal modeling given inadequate sample size for inference. This analysis was designated exploratory a priori given the small sample size and the distinct biological question it addresses relative to the primary SCCS.

## RESULTS

### Primary Analytic Cohort: NIH All of Us SCCS

#### Cohort Characteristics

From 633,547 enrolled participants in the All of Us Controlled Tier, 8,853 met SCCS eligibility criteria (≥90 days concurrent gabapentin–DHP-CCB use with documented discontinuation and ≥90 days follow-up). Median matched window: 180 days. Age ≥65: 3,497 (39.5%). CKD at baseline: 1,692 (19.1%). Discontinuation: stopped both (N=5,753, 65.0%), stopped gabapentin only (N=1,657), stopped CCB only (N=1,443).

#### Primary SCCS Findings

Cognitive symptom rates were significantly higher during combination use than after discontinuation (RR 1.26, 95% CI 1.19–1.33, p<0.001), as was the composite cognitive endpoint (RR 1.42, 95% CI 1.30–1.54, p<0.001) (Table 2). The cognitive signal was concentrated in patients who stopped both medications (N=5,753): cognitive symptoms RR 1.45 (95% CI 1.35–1.57) and composite cognitive endpoint RR 1.58 (95% CI 1.41–1.78), both p<0.001 — consistent with the largest cognitive benefit accruing to patients who discontinued both drugs. Secondary outcomes (falls, encephalopathy, new dementia) were all elevated during combination use and directionally concordant with the cognitive findings, with the stopped-both falls estimate similarly amplified (Table 2).

#### DHP-Class Specificity (Negative Control)

The SCCS was repeated in 2,011 patients co-prescribed gabapentin and a non-DHP CCB (verapamil or diltiazem; Table 3). Cognitive symptoms (RR 1.06, NS), falls (RR 1.20, NS), and encephalopathy (RR 1.19, NS) were null in the non-DHP arm, and the stopped-both cognitive rate ratio was 0.99 (NS). A nominally significant composite signal in the non-DHP arm (RR 1.28, p=0.009) was driven by new dementia and did not replicate in the stopped-both subgroup; we discuss this further below.

#### CKD Stratification

CKD amplified effects in both the primary and stopped-both analyses (Table 4). Cognitive symptom rate ratios were larger in CKD patients (1.45, 95% CI 1.29–1.62; N=1,692) than in non-CKD patients (1.20, 1.12–1.28; N=7,161), and the CKD encephalopathy rate ratio was markedly elevated (4.75, 1.62–13.96, p=0.003) on small event counts (PRE=19, POST=4). The stopped-both CKD amplification was preserved (cognitive symptom RR 1.68 in CKD vs 1.39 in non-CKD).

### Single-Center Triangulation Cohort: Rutgers CRDW SCCS

#### Cohort Characteristics

From 23,557 patients with documented concomitant gabapentin and DHP-CCB use, 2,404 met all eligibility criteria under the corrected medication-ascertainment approach (Table 1). The cohort was 67.5% age ≥65 (N=1,623) and 61.6% female (N=1,482); 22.1% (N=531) had a CKD diagnosis at baseline. Median matched window was 179 days (interquartile range 124–180), and median combination duration was 400 days (IQR 216–649). Discontinuation patterns: stopped both medications 45.0% (N=1,081), stopped gabapentin only 31.7% (N=763), stopped CCB only 23.3% (N=560).

**Table 1.** Cohort Characteristics — All of Us and CRDW Self-Controlled Case Series Cohorts.

| Characteristic | All of Us (N=8,853) | CRDW (N=2,404) |
| --- | --- | --- |
| ***Demographics*** |  |  |
| Age ≥65, N (%) | 3,497 (39.5%) | 1,623 (67.5%) |
| Female, N (%) | [TO FILL] | 1,482 (61.6%) |
| CKD at baseline, N (%) | 1,692 (19.1%) | 531 (22.1%) |
| Matched window, median (IQR), days | 180 | 179 (124–180) |
| Combination duration, median | [TO FILL] | 400 (216–649) |
| (IQR), days |  |  |
| **Discontinuation Pattern** |  |  |
| Stopped both medications, N (%) | 5,753 (65.0%) | 1,081 (45.0%) |
| Stopped gabapentin only, N (%) | 1,657 (18.7%) | 763 (31.7%) |
| Stopped CCB only, N (%) | 1,443 (16.3%) | 560 (23.3%) |

Cohort characteristics for All of Us primary analytic cohort (N=8,853) and CRDW single-center triangulation cohort (N=2,404) under corrected medication ascertainment. AoU values marked [TO FILL] pending extraction.

**Table 2.** All of Us SCCS — Primary Outcomes (N=8,853)

| Outcome | PRE Events | POST Events | RR (95% CI) | P-value | CRDW Reference |
| --- | --- | --- | --- | --- | --- |
| **Full Cohort (N=8,853)** |  |  |  |  |  |
| Falls | 537 | 350 | **1.53 (1.34–1.76)** | <0.001 | 0.94 (NS) |
| Cognitive symptoms | 2,675 | 2,127 | **1.26 (1.19–1.33)** | <0.001 | 1.19 (NS) |
| Composite cognitive† | 1,158 | 815 | **1.42 (1.30–1.54)** | <0.001 | 1.02 (NS) |
| Encephalopathy | 279 | 201 | **1.39 (1.20–1.61)** | <0.001 | NS |
| New dementia | 810 | 598 | **1.35 (1.22–1.51)** | <0.001 | — |
| **Stopped-Both Subgroup (N=5,753)** |  |  |  |  |  |
| Falls | 324 | 195 | **1.66 (1.39–1.98)** | <0.001 | 1.04 (NS) |
| Cognitive symptoms | 1,565 | 1,076 | **1.45 (1.35–1.57)** | <0.001 | 1.46 (1.09–1.94)* |
| Composite cognitive† | 712 | 450 | **1.58 (1.41–1.78)** | <0.001 | —‡ |

All of Us Controlled Tier Dataset C2024Q3R9. PRE = final matched window of concurrent use; POST = equal-duration window following discontinuation. †Composite = cognitive symptoms, encephalopathy, delirium, or new dementia. CRDW Reference column shows corrected single-center estimates (N=2,404; Tables 5–6). \* $p < 0.05$ ; ‡ composite not separately reported for CRDW stopped-both. NS = not significant.

**Table 3.** Mechanistic Specificity — DHP-CCB vs Non-DHP CCB (All of Us Controlled Tier)

| Outcome | DHP-CCB (N=8,853) | Non-DHP CCB (N=2,011) |
| --- | --- | --- |
| Falls | **1.53 (1.34–1.76)\*\*** | **1.20 (0.95–1.51) NS** |
| Cognitive symptoms | **1.26 (1.19–1.33)\*\*** | **1.06 (0.94–1.19) NS** |
| Composite cognitive† | **1.42 (1.30–1.54)\*\*** | **1.28 (1.07–1.54)\*\*** |
| Encephalopathy | **1.39 (1.16–1.66)\*\*** | **1.19 (0.74–1.92) NS** |
| *Stopped-Both Cognitive Symptoms* | **1.45 (1.35–1.57)\*\*** | **0.99 (0.84–1.17) NS** |
| *Stopped-Both Falls* | **1.66 (1.39–1.98)\*\*** | **1.54 (1.16–2.05)\*\*** |

*DHP-CCB: gabapentin + amlodipine/felodipine/nifedipine (N=8,853). Non-DHP CCB: gabapentin + verapamil/diltiazem (N=2,011). [Stopped Both] rows = stopped-both subgroup only. \*\* p<0.01; NS = not significant.*

**Table 4.** All of Us CKD-Stratified SCCS (N=8,853)

| Outcome | PRE | POST | RR (95% CI) | P-value | CRDW Reference (corrected) |
| --- | --- | --- | --- | --- | --- |
| ***Non-CKD (N=7,161)*** |  |  |  |  |  |
| Falls | 403 | 273 | **1.48 (1.27–1.72)** | <0.001 | 0.87 |
| Cognitive symptoms | 1,926 | 1,609 | **1.20 (1.12–1.28)** | <0.001 | 1.14 |
| *Stopped-Both Cognitive Symptoms* | 1,162 | 836 | **1.39 (1.27–1.52)** | <0.001 | 1.39 |
| ***CKD (N=1,692)*** |  |  |  |  |  |
| Falls | 134 | 77 | **1.74 (1.31–2.30)** | <0.001 | 1.29 |
| Cognitive symptoms | 749 | 518 | **1.45 (1.29–1.62)** | <0.001 | 1.28 |
| Encephalopathy | 19 | 4 | **4.75 (1.62–13.96)** | 0.003 | — |
| *Stopped-Both Cognitive Symptoms* | 403 | 240 | **1.68 (1.43–1.97)** | <0.001 | 1.60 |

*CKD defined as any CKD diagnosis on or before combination start date. CRDW Reference column shows corrected single-center point estimates (N=2,404). Encephalopathy in CKD based on small event counts (PRE=19, POST=4) — interpret with caution.*

#### Primary SCCS Analysis

At the primary lag-0 specification, the corrected CRDW analysis showed concordant direction of effect with point estimates closer to the null (Table 5). Cognitive symptoms RR was 1.19 (95% CI 0.97–1.45), composite cognitive endpoint RR was 1.02 (0.83–1.25), falls RR was 0.94 (0.77–1.15), and encephalopathy did not differ between PRE and POST. The single-center cohort (N=2,404) is smaller than All of Us and correspondingly less powered.

**Table 5.**
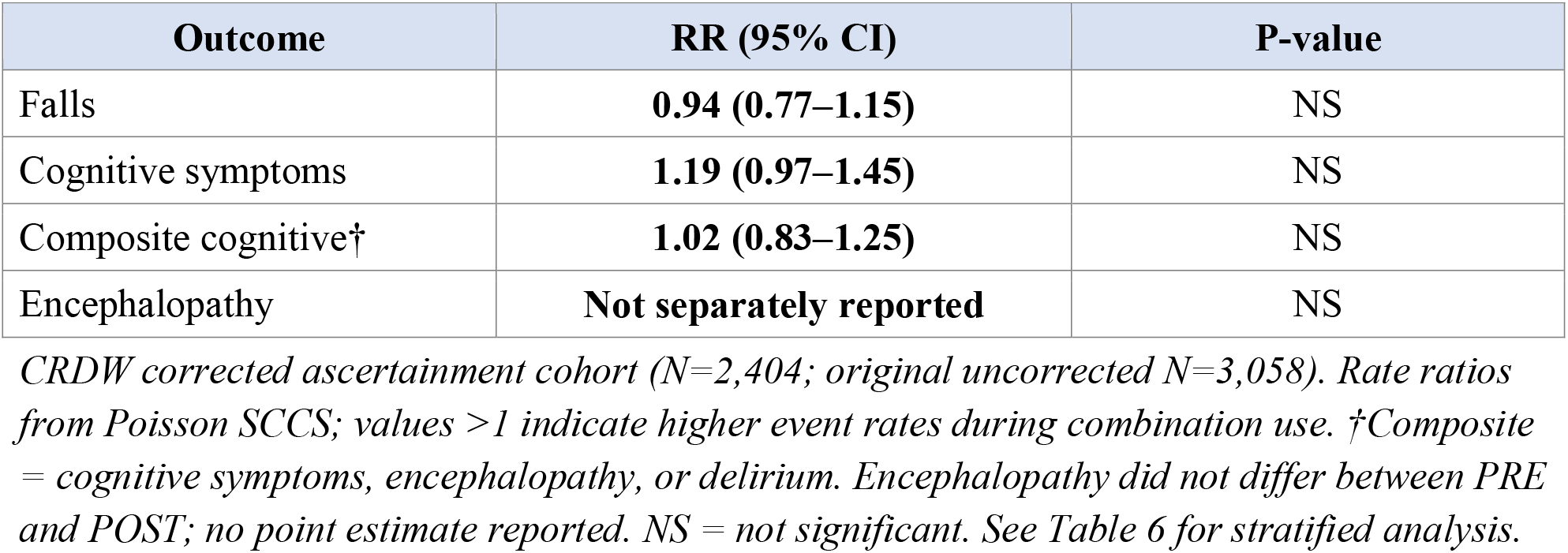
Single-Center CRDW SCCS — Primary Outcomes Under Corrected Medication Ascertainment (N=2,404)

| Outcome | RR (95% CI) | P-value |
| --- | --- | --- |
| Falls | **0.94 (0.77–1.15)** | NS |
| Cognitive symptoms | **1.19 (0.97–1.45)** | NS |
| Composite cognitive† | **1.02 (0.83–1.25)** | NS |
| Encephalopathy | **Not separately reported** | NS |

#### Stratified Analysis by Discontinuation Pattern

Within the CRDW cohort, stratification by discontinuation pattern identified one signal that reached statistical significance: cognitive symptoms in patients who discontinued both medications simultaneously (RR 1.46, 95% CI 1.09–1.94, p<0.05; Table 6). Stopped-both falls (RR 1.04, NS) and stopped-both composite outcomes did not differ significantly between PRE and POST periods, and stopping either drug alone produced non-significant point estimates for all outcomes.

**Table 6.** CRDW Stratified Analysis by Discontinuation Pattern and Lag-Extended Stopped-Both Cognitive Symptoms.

| Discontinuation Type | N | Falls RR (95% CI) | Cognitive Symptoms RR (95% CI) |
| --- | --- | --- | --- |
| **Stopped both medications** | 1,081 | 1.04 (NS) | **1.46 (1.09–1.94)\*** |
| Stopped gabapentin only | 763 | 1.18 (0.81–1.71) | 1.09 (0.73–1.63) |
| Stopped CCB only | 560 | **0.65 (0.45–0.95)\*†** | 0.87 (0.59–1.29) |
| **Stopped both × Age ≥65** | 738 | 0.96 (0.68–1.35) | **1.74 (1.22–2.50)\*\*** |

| Lag Window | N | Stopped-Both Cognitive Symptoms RR (95% CI) | P-value |
| --- | --- | --- | --- |
| Lag-0 (primary) | 1,081 | **1.46 (1.09–1.94)** | 0.012 |
| 30-day lag | 836 | **1.56 (1.12–2.18)** | 0.010 |
| 60-day lag | 679 | **1.69 (1.15–2.48)** | 0.008 |

Panel B. Protopathic-bias sensitivity analysis of the stopped-both cognitive symptoms signal in the CRDW corrected-ascertainment cohort. POST window shifted forward by 30 and 60 days to evaluate lag sensitivity.

#### Age ≥65 Stopped-Both Subgroup

Within the older patient subgroup (age ≥65, N=1,623; the population most clinically relevant to deprescribing decisions), the stopped-both cognitive symptoms signal was strengthened: PRE = 82 events, POST = 47 events, RR = 1.74 (95% CI 1.22–2.50, p=0.003). The age ≥65 stopped-both falls signal was null (RR 0.96, 0.68–1.35).

#### Single-Drug Discontinuation Subgroups

Among patients who discontinued only the gabapentinoid (N=763), point estimates were directionally consistent but did not reach significance (cognitive symptoms RR 1.09, 95% CI 0.73–1.63; falls RR 1.18, 0.81–1.71). Among patients who discontinued only the DHP-CCB (N=560), the cognitive symptoms point estimate was null (RR 0.87, 0.59–1.29), while the falls outcome showed an inverse signal (RR 0.65, 0.45–0.95, p=0.031). The inverse falls signal is most parsimoniously explained by discontinuation-decision protopathic bias (Discussion); cognitive symptoms were not directionally inverse in any subgroup.

#### Sex-Stratified Sensitivity Analysis

A sex-stratified sensitivity analysis showed that the female stratum (N=1,482) had a significantly elevated cognitive symptom rate during combination use at the primary lag-0 specification, even without restricting to the stopped-both subgroup: RR 1.32 (95% CI 1.02–1.71, p=0.037). Although this single-stratum result should be interpreted as exploratory and was not pre-specified, it is consistent with the overall directional pattern and provides further support for the cognitive-symptom-specific reversibility signal as the most robust finding in the corrected single-center cohort.

#### Cross-Cohort CKD-Amplification Pattern

The corrected CRDW analysis preserved the CKD-amplification directional pattern observed in All of Us across all three primary outcomes (CRDW CKD vs non-CKD: falls 1.29 vs 0.87, cognitive symptoms 1.28 vs 1.14, stopped-both cognitive symptoms 1.60 vs 1.39). Although individual stratum-specific results did not reach conventional significance in the smaller corrected single-center cohort, the consistent CKD>non-CKD ordering across outcomes is concordant with the All of Us pattern.

#### Lag-Period Sensitivity Analysis

The stopped-both cognitive symptoms signal strengthened monotonically across protopathic-bias lag conditions: rate ratio 1.46 (95% CI 1.09–1.94, p=0.012) at lag-0, 1.56 (1.12–2.18, p=0.010) at 30-day lag (N=836), and 1.69 (1.15–2.48, p=0.008) at 60-day lag (N=679). This is the opposite of the pattern protopathic bias would produce, and is consistent with pharmacological washout, with the 30- and 60-day windows broadly aligned with the half-lives of the index drugs (gabapentin 5–7 hours under normal renal function; amlodipine 30–50 hours).

### Cross-Cohort Agreement on Stopped-Both Cognitive Symptoms

The stopped-both cognitive symptoms finding shows close concordance between the two independent datasets, with All of Us and CRDW point estimates of 1.45 and 1.46, respectively, and the CRDW estimate strengthening monotonically across 0-, 30-, and 60-day lag windows (Table 6 Panel B). We consider this cross-cohort convergence on the stopped-both cognitive symptoms outcome the principal triangulation evidence supporting reversibility upon dual discontinuation; the larger All of Us cohort provides the primary inferential basis, with CRDW providing single-center reproducibility under corrected medication ascertainment.

### Established Dementia Cohort (Supplementary Material)

Results of the exploratory established dementia cohort analysis are reported in the Supplementary Material. Under corrected medication ascertainment, the analytic cohort comprised 246 patients (101 DHP-CCB-exposed; 145 no-CCB), and the IPTW-adjusted hazard ratio for incident encephalopathy in DHP-CCB users versus no-CCB users was 0.73 (95% CI 0.30–1.76, p=0.48). The corrected null finding contrasts with our preliminary analysis under uncorrected ascertainment, in which intravenous nicardipine prescriptions during inpatient hypertensive emergencies had been classified as DHP-CCB exposure. This exploratory result does not refute the primary cross-cohort SCCS findings, which apply to a distinct population without prior dementia.

## DISCUSSION

Cognitive symptom rates declined following discontinuation of concomitant gabapentin–DHP-CCB therapy across two independent datasets (All of Us N=8,853, CRDW N=2,404), with the largest declines among patients who stopped both medications and closely concordant stopped-both estimates between cohorts (All of Us RR 1.45; CRDW RR 1.46). For clinicians caring for older adults presenting with cognitive complaints while taking this combination, these findings support structured medication review and, where clinically appropriate, deprescribing of one or both agents — with the expectation that some portion of the cognitive burden may be reversible. The DHP-class specificity of the cognitive signal (null with verapamil/diltiazem in All of Us) and CKD amplification across cohorts point to the gabapentin–DHP-CCB combination, rather than antihypertensive discontinuation broadly, as the relevant exposure.

The clinical signal is consistent across cohorts and across primary outcomes. In All of Us, cognitive symptoms were 26% more frequent during combination use than after discontinuation, and 45% more frequent in patients who subsequently stopped both medications. The gradient by discontinuation type, with the largest reduction observed when both drugs are removed, is consistent with both medications contributing to the cognitive signal, though we cannot resolve their individual contributions from these data. Secondary outcomes (falls, encephalopathy) were directionally consistent in All of Us.

### External Replication and Cross-Cohort Triangulation

The All of Us replication addresses the most important alternative explanation: if the reversal signal reflected non-specific confounding by stopping any antihypertensive agent, it would appear with verapamil and diltiazem, which carry equivalent antihypertensive potency — but cognitive, fall, and encephalopathy outcomes were null in the non-DHP arm. The nominally significant non-DHP composite signal was driven by new dementia and did not replicate in the stopped-both subgroup; this may reflect residual confounding by cardiac disease severity in verapamil/diltiazem-treated patients. Effect sizes in All of Us are modestly smaller than in prior CRDW analyses, consistent with the healthy volunteer effect of the All of Us enrollment model.

### Discontinuation-Pattern Stratification

The stratified Table 6 Panel A results help interpret which discontinuation pattern carries the cognitive signal. The reversal signal was strongest in patients who discontinued both medications (RR 1.46) and was directionally consistent (though underpowered) in the stopped-gabapentinoid-only subgroup. Stopping only the DHP-CCB while continuing the gabapentinoid did not produce a cognitive benefit. The inverse falls signal in the stopped-CCB-only subgroup is most parsimoniously explained by discontinuation-decision protopathic bias (limitation 6).

### Protopathic Bias

The lag analyses argue against protopathic bias as the principal explanation: the stopped-both cognitive symptoms signal strengthened rather than attenuated as the immediate post-discontinuation period was excluded, with the same directional pattern in both cohorts. While not definitive, this raises the evidentiary bar for an artifactual interpretation.

### Biological Plausibility

The pattern observed — reversibility upon discontinuation, DHP-class specificity, and CKD amplification — is broadly compatible with a shared pharmacological action involving neuronal calcium signaling, though observational data cannot establish mechanism directly.^4–8^ The CKD-amplification gradient is consistent with gabapentin’s renal clearance, though greater baseline susceptibility among patients with CKD cannot be excluded.

### Strengths and Limitations

The self-controlled case series design eliminates confounding by all time-invariant characteristics, and duration matching ensures identical person-time. The DHP-subtype specificity control and cross-cohort triangulation in two independent datasets provide independent corroboration, with the close numerical concordance of the stopped-both cognitive symptoms estimate across cohorts (1.45 vs 1.46) unlikely to occur by chance under the null.

Several limitations warrant discussion. First, we relied on EHR prescription records to ascertain chronic outpatient medication exposure; residual misclassification (non-adherence, out-of-system prescriptions) remains possible. Second, the CRDW triangulation cohort was drawn from a single academic health system in northern New Jersey, and the All of Us Controlled Tier mitigates this at scale but is itself subject to volunteer-cohort selection effects. Replication in Medicare fee-for-service data via the IFH Data Core (DUA RSCH-2024-70249) is in progress.

Third, our medication-ascertainment approach (excluding discontinued orders and intravenous formulations) is a refinement over an earlier extraction pipeline; estimates are correspondingly more conservative.

Fourth, the single-center CRDW analysis was sensitive to ascertainment specification: under the corrected approach, three primary CRDW outcomes did not reach significance, while the stopped-both cognitive symptoms signal was preserved.

Fifth, the self-controlled design cannot account for time-varying confounders that change between PRE and POST windows, and EHR-based outcome ascertainment may undercount true event rates and differentially capture events during active healthcare engagement. The All of Us composite outcome showed a nominally significant result in the non-DHP arm, warranting cautious interpretation.

Sixth, the matched-window SCCS design provides only partial protection against discontinuation-decision protopathic bias, which can be substantial when discontinuation is itself event-triggered. The inverse falls signal in the stopped-CCB-only subgroup (RR 0.65, 95% CI 0.45–0.95, p=0.031) likely reflects this; the lag analyses partially address it for cognitive symptoms but cannot fully eliminate event-driven discontinuation as a confounding mechanism for falls.

## CONCLUSIONS

Cognitive symptoms associated with concomitant gabapentin–DHP-CCB use declined after discontinuation in two independent cohorts, with the largest reductions among patients who stopped both medications (All of Us RR 1.45; CRDW RR 1.46; 1.74 in adults aged ≥65). These findings identify cognitive symptoms in older adults taking this combination as a potentially reversible, medication-related contribution to clinical presentation, and support structured medication review and deprescribing assessment when clinicians encounter cognitive complaints in patients on gabapentin and a DHP-CCB. Secondary outcomes were directionally concordant in the All of Us cohort. External replication — including planned analyses in Medicare fee-for-service data — and prospective evaluation of structured deprescribing in this population are the highest-value next steps. An exploratory analysis in patients with established dementia is reported in the Supplementary Material.

## Data Availability

The Rutgers Clinical Research Data Warehouse (CRDW) data that support the findings of this study are available through the Rutgers University health system subject to institutional data governance review. NIH All of Us Controlled Tier data are available to credentialed researchers through the All of Us Researcher Workbench (https://workbench.researchallofus.org). Analytic code is available from the corresponding author upon reasonable request.

## ACKNOWLEDGMENTS

Data used in this research were obtained from the Clinical Research Data Warehouse (CRDW), a joint initiative of RWJBarnabas Health and Rutgers, The State University of New Jersey, and are used with permission of the Data Governance Council. Data were accessed through the Rutgers Clinical Research Data Warehouse and the NIH All of Us Research Program Controlled Tier Dataset (C2024Q3R9). We gratefully acknowledge All of Us participants for their contributions, without whom this research would not have been possible. We also thank the National Institutes of Health’s All of Us Research Program for making available the participant data examined in this study.

## AUTHOR CONTRIBUTIONS

James W. Green: Conceptualization, Methodology, Formal Analysis, Data Curation, Writing – Original Draft, Visualization. Joshua Kaplan: Conceptualization, Writing – Review & Editing. Branimir Ljubic: Resources, Data Curation, Writing – Review & Editing. Michael Schulewski: Resources, Data Curation, Writing – Review & Editing. Suril Gohel: Supervision, Writing – Review & Editing. Sharon Sanz Simon: Writing – Review & Editing. Luciana Mascarenhas Fonseca: Writing – Review & Editing. Michal Schnaider Beeri: Writing – Review & Editing. Laura Byham-Gray: Supervision, Writing – Review & Editing. Barbara Tafuto: Supervision, Writing – Review & Editing.

## ETHICS STATEMENT

This study used de-identified electronic health record data from the Rutgers Clinical Research Data Warehouse (CRDW) and the NIH All of Us Research Program Controlled Tier. Both data sources were determined to constitute non-human subjects research per 45 CFR 46.102(d). The CRDW analysis was conducted under the Rutgers Data Governance Council’s data use agreement; the All of Us analysis was conducted under the All of Us IRB-approved protocol.

Informed consent was not required.

## DATA AVAILABILITY STATEMENT

CRDW data are available through Rutgers with IRB approval and Data Governance Council permission. All of Us data are available to credentialed researchers through the Researcher Workbench (https://workbench.researchallofus.org).

## SUPPLEMENTARY MATERIAL

***Reversibility of Gabapentin–DHP-CCB-Associated Cognitive Outcomes Following Discontinuation: A Cross-Cohort Self-Controlled Case Series in the All of Us Research Program with Single-Center CRDW Triangulation***

### Overview

This Supplementary Material reports an exploratory analysis of incident encephalopathy in patients with established dementia who initiated gabapentin while receiving a calcium channel blocker (CCB). Under corrected medication ascertainment, the analysis was null. The analysis is reported separately from the main manuscript because (1) it addresses a different biological question—incident encephalopathy in an already-demented population, rather than reversibility of cognitive and falls outcomes upon discontinuation; (2) the analytic cohort under corrected ascertainment is small (N=246) and drawn from a single academic health system; and (3) the non-DHP CCB descriptive arm under corrected ascertainment dropped to N=8, below the threshold for inferential analysis. Findings should be considered hypothesis-generating and do not refute the primary cross-cohort SCCS reversibility findings, which apply to a distinct population (patients without prior dementia).

### Methods: Established Dementia Cohort

#### Study Population

An exploratory analysis was conducted in patients within the Rutgers Clinical Research Data Warehouse (CRDW; 2018–2024) with prior dementia, defined as any documented dementia diagnosis (ICD-10 F01–F03, G30, G31.0, G31.1, G31.83) recorded ≥1 day before gabapentinoid initiation, who initiated a gabapentinoid (gabapentin or pregabalin) and had ≥30 days of subsequent follow-up. The supplementary analysis uses a broader exposure definition than the primary SCCS (gabapentinoid versus gabapentin only) to maximize statistical power in this small subpopulation. Under corrected medication ascertainment—excluding discontinued orders and intravenous formulations, as described in the main manuscript Methods (Medication Exposure Ascertainment)—eligible patients were classified by DHP-CCB exposure status at gabapentinoid index (any DHP-CCB prescription on or before the index date). The analytic cohort comprised 246 patients: 101 DHP-CCB-exposed (amlodipine, nifedipine, felodipine, or nicardipine restricted to chronic outpatient oral therapy) and 145 no-CCB. The non-DHP CCB descriptive arm under corrected ascertainment contained 8 patients (verapamil or diltiazem) with 0 encephalopathy events and is not reported as a separate stratum, given that DHP-subtype specificity in this research program is established by the All of Us non-DHP CCB negative control analysis (N=2,011) reported in the main manuscript.

#### Outcome and Statistical Analysis

The primary outcome was incident encephalopathy after gabapentinoid initiation, defined by ICD-10 codes G92, G93.4, G93.40, G93.41, and G93.49. An inverse-probability-of-treatment-weighted (IPTW) Cox proportional hazards model was fitted comparing DHP-CCB-exposed and no-CCB strata, adjusting for age, sex, baseline CKD, dementia duration before gabapentinoid index, and gabapentinoid arm (gabapentin vs pregabalin). Propensity scores for DHP-CCB exposure were estimated using logistic regression on the same covariates; IPTW weights were truncated at the 1st and 99th percentiles. Falls, delirium, and dementia-medication-initiation outcomes from the prior version of this supplementary analysis were not re-analyzed under corrected ascertainment because the corrected analytic cohort is materially different in composition; consequently, only the corrected encephalopathy outcome is reported. Selectively re-running some outcomes but not others would be inconsistent.

#### Note on Corrected Medication Ascertainment

This analysis applies the corrected medication-ascertainment approach described in the main manuscript Methods, excluding prescription records with ACTIVE_ORDER_STATUS = ‘Discontinued Medication’ and intravenous formulations identified by route or product-name pattern.

Earlier preliminary versions of this supplementary analysis included inpatient intravenous nicardipine prescriptions—captured during inpatient hypertensive emergencies—in the DHP-CCB-exposed arm, which inflated the DHP-CCB arm size and produced a preliminary positive encephalopathy signal that did not survive correction.

Restricting ascertainment to chronic outpatient oral DHP-CCB use is consistent with the broader hypothesis articulated in the main manuscript that the gabapentin–DHP-CCB pharmacodynamic interaction operates through sustained chronic exposure in cognitively-intact populations, rather than through acute toxicity in patients with established dementia.

#### Results: Established Dementia Cohort

Under corrected medication ascertainment, the IPTW-adjusted hazard ratio for incident encephalopathy in DHP-CCB users (N=101) versus no-CCB users (N=145) was 0.73 (95% CI 0.30–1.76, p=0.48; eTable S1). The corrected null finding is consistent with the broader interpretation articulated in the main manuscript: that the gabapentin–DHP-CCB pharmacodynamic interaction operates through sustained chronic outpatient exposure in cognitively-intact populations, rather than through acute toxicity in patients with established dementia.

DHP-class specificity in this exploratory cohort cannot be formally tested because the non-DHP CCB descriptive arm (N=8) is below the threshold for inferential analysis. Mechanistic specificity for DHP-class blockade is addressed in the main manuscript via the All of Us non-DHP CCB negative control (N=2,011), which showed null results across falls, cognitive symptoms, and encephalopathy in the non-DHP arm in direct contrast to the highly significant signals in the DHP arm. The present exploratory finding does not refute the primary cross-cohort SCCS reversibility findings, which apply to a distinct population (patients without prior dementia).

## Supplementary Table

**eTable S1.** Established Dementia Cohort — Encephalopathy IPTW Cox Analysis Under Corrected Medication Ascertainment (N=246).

| Outcome | DHP-CCB<br>(N=101) | No CCB<br>(N=145) | IPTW HR (95% CI)<br>vs No CCB | P-value |
| --- | --- | --- | --- | --- |
| Incident<br>encephalopathy | *(exposed)* | *reference* | **0.73 (0.30–1.76)** | 0.48 |

*Cox proportional hazards model with inverse probability of treatment weighting (IPTW); reference group = no-CCB users. Cohort restricted to patients with documented prior dementia (ICD-10 F01–F03, G30, G31.0, G31.1, G31.83) recorded $\geq 1$ day before gabapentinoid (gabapentin or pregabalin) initiation, with $\geq 30$ days subsequent follow-up. Encephalopathy defined by ICD-10 G92, G93.4, G93.40, G93.41, or G93.49. IPTW Cox model adjusted for age, sex, baseline CKD, dementia duration before gabapentinoid index, and gabapentinoid arm. Non-DHP CCB descriptive arm (N=8) excluded from formal modeling and reported in the Methods only; zero encephalopathy events were observed in that small descriptive arm. Abbreviations: HR, hazard ratio; IPTW, inverse probability of treatment weighting.*

