## Supplemental Material for "Do Cognitive Symptoms Improve When Gabapentin and Dihydropyridine Calcium Channel Blocker Therapy Are Stopped? A Self-Controlled Case Series" (from another version of this preprint: version 1, 2026-04-01)

MP2 — Reversibility of Cognitive Effects Associated with Concomitant Gabapentin and Dihydropyridine Calcium Channel Blocker Use

MP3 — Sequence-Dependent Amplification of Gabapentinoid-Associated Dementia Risk by Dihydropyridine Calcium Channel Blockers

#### Supplementary Methods

##### MP3: Propensity Score Estimation and IPTW Diagnostics

Propensity scores were estimated using L2-penalized logistic regression (C=1.0; C-statistic 0.560). Stabilized inverse probability of treatment weights (IPTW) used the average treatment effect in the treated (ATT) estimand, truncated at the 1st and 99th percentiles (bounds: 0.799–1.275). The effective sample size after weighting was 4,429 (99.5% of N=4,451), indicating minimal information loss. Full IPTW diagnostics including propensity score overlap, weight distribution, and truncation sensitivity are presented in eFigure S2. Post-weighting covariate balance for Population 4 is presented in eTable S1 (maximum |SMD| = 0.097). The E-value for the primary HR of 2.23 is 3.89; the E-value for the lower confidence bound (HR 1.43) is 2.21, indicating that an unmeasured confounder would need at least a 3.9-fold association with both treatment selection and dementia to fully explain the observed effect.

##### MP2: Sensitivity Analysis and E-Values

The E-value for the SCCS cognitive symptoms rate ratio of 1.67 is 2.73. The E-value for the dementia acceleration cohort encephalopathy hazard ratio of 3.18 is 5.81 (lower confidence bound E-value: 2.06). The dementia acceleration cohort Cox model was unadjusted in the primary analysis given sparse events (N=320); an adjusted sensitivity analysis incorporating age, sex, and CKD status is presented in eTable S4.

### eFigure S2. Population 4 IPTW Diagnostics — Gabapentin vs Pregabalin (Chronic DHP-CCB Users)

**Population 4 IPTW Diagnostics — Gabapentin vs Pregabalin (Chronic DHP-CCB Users)**

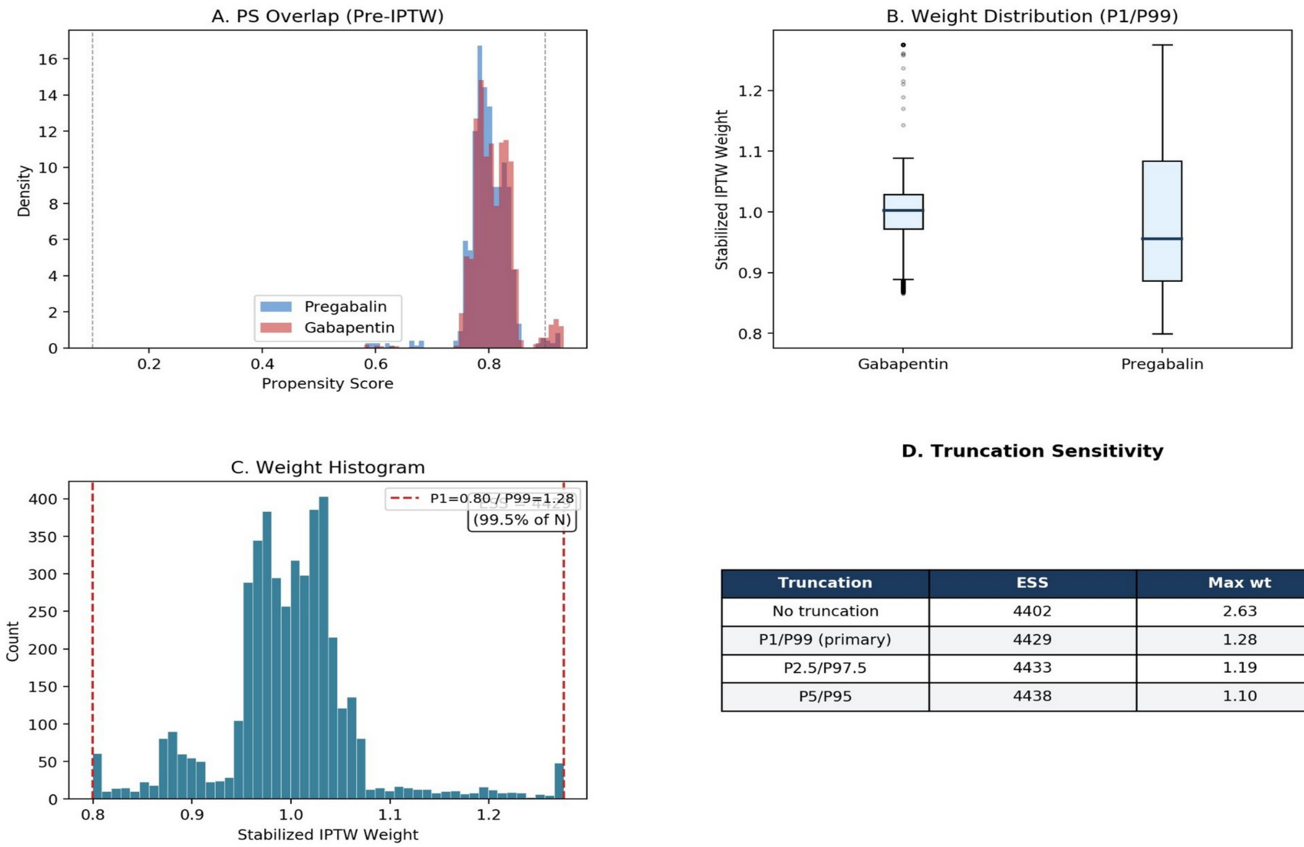


**D. Truncation Sensitivity**

| Truncation | ESS | Max wt |
|---|---|---|
| No truncation | 4402 | 2.63 |
| P1/P99 (primary) | 4429 | 1.28 |
| P2.5/P97.5 | 4433 | 1.19 |
| P5/P95 | 4438 | 1.10 |

Panel A: Propensity score overlap pre-IPTW showing adequate positivity across both treatment arms. Panel B: Stabilized IPTW weight distribution by treatment arm. Panel C: Weight histogram with P1/P99 truncation bounds (red dashed lines; 99.5% of sample retained). Panel D: Truncation sensitivity — effective sample size and maximum weight are stable across P1/P99, P2.5/P97.5, and P5/P95 thresholds, confirming robustness of the primary truncation choice. PS = propensity score; IPTW = inverse probability of treatment weighting; ESS = effective sample size.

### eTable S1. Covariate Balance Before and After IPTW — Population 4 (MP3)

| Covariate | Gaba (post) | Gaba (pre) | Pregab (post) | Pregab (pre) | \|SMD\| post | \|SMD\| pre | Balanced |
|---|---|---|---|---|---|---|---|
| on_non_dhp | 0.043 | 0.049 | 0.025 | 0.020 | 0.097 | 0.160 | YES |
| has_mci | 0.010 | 0.008 | 0.015 | 0.019 | 0.049 | 0.094 | YES |
| on_beta | 0.430 | 0.445 | 0.419 | 0.370 | 0.023 | 0.153 | YES |
| ccb_duration | 795.3 | 792.6 | 801.0 | 805.7 | 0.018 | 0.043 | YES |
| sex_male | 0.468 | 0.475 | 0.461 | 0.441 | 0.014 | 0.069 | YES |
| has_ckd | 0.256 | 0.262 | 0.252 | 0.234 | 0.011 | 0.064 | YES |
| on_dem_med_bl | 0.014 | 0.014 | 0.015 | 0.013 | 0.007 | 0.010 | YES |
| has_fall_hx | 0.073 | 0.072 | 0.074 | 0.077 | 0.005 | 0.017 | YES |
| on_hctz | 0.207 | 0.207 | 0.209 | 0.205 | 0.005 | 0.005 | YES |
| age | 69.58 | 69.66 | 69.55 | 69.28 | 0.003 | 0.031 | YES |
| age_ge65 | 0.684 | 0.689 | 0.683 | 0.662 | 0.003 | 0.058 | YES |
| on_ace_arb | 0.469 | 0.473 | 0.468 | 0.454 | 0.002 | 0.039 | YES |

All 12 covariates achieved balance after IPTW (|SMD| < 0.10). Maximum post-weighting |SMD| = 0.097 (on_non_dhp). Comorbidities beyond CKD were not included as IPTW model covariates given the focused propensity score design; descriptive prevalence of additional comorbidities is provided in Table 1 of the main manuscript. Gaba = gabapentin initiators; Pregab = pregabalin initiators; post = after IPTW; pre = before IPTW. on_non_dhp = non-DHP CCB use; has_mci = mild cognitive impairment at baseline; on_beta = beta-blocker use; ccb_duration = days of prior CCB therapy; has_ckd = chronic kidney disease at baseline; on_dem_med_bl = dementia medication at baseline; has_fall_hx = fall history; on_hctz = thiazide diuretic use; on_ace_arb = ACE inhibitor or ARB use.

### eTable S2. Falsification Outcome Analysis — Population 4 (MP3)

| Falsification Outcome | Events | OR | 95% CI | Null? |
|---|---|---|---|---|
| Appendicitis | 60 | 1.25 | 0.59–2.64 | YES |
| Herpes zoster | 672 | 1.39 | 1.10–1.76 | NO |
| Skin cancer | 305 | 0.84 | 0.63–1.12 | YES |
| Cataract | 624 | 0.84 | 0.69–1.03 | YES |
| Osteoarthritis | 2,568 | 0.93 | 0.84–1.04 | YES |
| GERD | 2,700 | 0.91 | 0.82–1.01 | YES |
| UTI | 2,547 | 1.06 | 0.95–1.18 | YES |

Falsification outcomes were pre-specified conditions with no biologically plausible link to the gabapentin-CCB combination. Six of 7 outcomes were null (OR crossing 1.0). Herpes zoster showed a nominally elevated OR (1.39, 95% CI 1.10–1.76), likely reflecting the older, multimorbid nature of the CCB+gabapentin population rather than a causal effect. OR = odds ratio; CI = confidence interval.

#### eTable S3. E-Value Summary — MP1, MP2, and MP3 Primary Estimates

| Analysis | HR/RR | CI Lower | E-value | E-value (CI lb) |
|---|---|---|---|---|
| MP3 Pop 4 primary | 2.23 | 1.43 | 3.89 | 2.21 |
| MP3 broader CCB-first | 2.48 | 1.55 | 4.40 | 2.47 |
| MP3 recency 180d | 3.97 | 1.38 | 7.40 | 2.10 |
| MP3 acceleration encephalopathy | 2.09 | 1.19 | 3.60 | 2.10 |
| MP2 SCCS cognitive symptoms | 1.67 (RR) | 1.38 | 2.73 | 2.10 |
| MP2 SCCS falls | 1.34 (RR) | 1.11 | 2.01 | 1.46 |
| MP2 acceleration encephalopathy | 3.18 | 1.36 | 5.81 | 2.06 |
| MP1 interaction HR | 2.22 | 1.42 | 3.87 | 2.19 |
| MP1 DHP-specific HR | 3.20 | 1.81 | 5.85 | 3.02 |

E-values indicate the minimum strength of association that an unmeasured confounder would need with both treatment and outcome to fully explain the observed effect. E-values are presented for the point estimate and for the lower bound of the 95% confidence interval. HR = hazard ratio; RR = rate ratio; CI lb = confidence interval lower bound. MP1 = CCB interaction paper (JAMA); MP2 = reversibility paper; MP3 = sequence-dependence paper.
